# Quantifying Contact Patterns in Response to COVID-19 Public Health Measures in Canada

**DOI:** 10.1101/2021.03.11.21253301

**Authors:** Gabrielle Brankston, Eric Merkley, David N. Fisman, Ashleigh R. Tuite, Zvonimir Poljak, Peter J. Loewen, Amy L. Greer

## Abstract

**Background:** A variety of public health measures have been implemented during the COVID-19 pandemic in Canada to reduce contact between individuals.

**Objective:** The objective of this study was to construct contact patterns to evaluate the degree to which social contacts rebounded to normal levels, as well as direct public health efforts toward age- and location-specific settings.

**Design:** Four population-based cross-sectional surveys.

**Setting:** Canada.

**Participants:** Members of a paid panel representative of Canadian adults by age, gender, official language, and region of residence.

**Methods:** Respondents provided information about the age and setting for each direct contact made in a 24-hour period. Contact matrices were constructed and contacts for those under the age of 18 years imputed. The next generation matrix approach was used to estimate the reproduction number (R_t_) for each survey. Respondents with children estimated the number of contacts their children made in school and extracurricular settings.

**Results:** Estimated R_t_ values were 0.49 (95% CI: 0.29-0.69) for May, 0.48 (95% CI: 0.29-0.68) for July, 1.06 (95% CI: 0.63-1.52) for September, and 0.81 (0.47-1.17) for December. The highest proportion of reported contacts occurred within the home (51.3% in May), in ‘other’ locations (49.2% in July) and at work (66.3% and 65.4% in September and December). Respondents with children reported an average of 22.7 (95% CI: 21.1-24.3) (September) and 19.0 (95% CI 17.7-20.4) (December) contacts at school per day per child in attendance.

**Conclusion:** The skewed distribution of reported contacts toward workplace settings in September and December combined with the number of reported school-related contacts suggest that these settings represent important opportunities for transmission emphasizing the need to ensure infection control procedures in both workplaces and schools.

## Introduction

In March, 2020, as transmission of SARS-CoV-2, the virus that causes Coronavirus Disease 2019 (COVID-19), was increasing across Canada, provincial and local governments implemented a variety of non-pharmaceutical public health measures (1). As a main route of transmission of SARS-CoV-2 is close contact with an infected individual (2), these restrictions were implemented to reduce contact between individuals and included a variety of physical distancing measures (1). Canadian modelling studies have estimated that a 45-60% reduction in direct, close proximity contacts would be sufficient to suppress community transmission, reduce the number of cases of COVID-19, and protect the health care system from becoming overwhelmed (3,4). However, these estimates were based on model assumptions regarding expected changes in contact patterns under physical distancing measures during the COVID-19 pandemic.

For directly transmitted respiratory pathogens, transmission opportunities exist anywhere individuals can have direct close proximity contact including households, workplaces, and schools. Quantifying age-specific contact patterns improves our understanding of disease transmission, allows for the estimation of important epidemic parameters such as the reproduction number (5,6), and provides empirical data for use in mathematical models which typically rely on estimates of contact patterns that have been collected in previous studies (6). Further, quantifying changes in contact patterns at different time points during a pandemic allows us to evaluate the effectiveness of public health measures, and identify the degree to which contacts have returned to pre-pandemic levels, as well as direct improved public health messaging efforts toward age- and location-specific settings.

The reproduction number can be estimated from the growth of cases in surveillance data (7,8) however, due to variations in testing and contact tracing as well as delays in reporting, case counts are not always a reliable indication of transmission. Real-time estimation of the effective reproduction number (R_t_) involves examining the ratio of change in the contact matrix from one generation of infection to the next (9). For respiratory infections that are transmitted by direct contact, we can use survey-derived contact matrices to estimate how changes in contact patterns (where each contact represents an opportunity for transmission) influence the reproduction number (5,6). If the average number of opportunities for transmission declines by a certain amount, so should the corresponding value of the reproduction number.

A number of recent studies have examined the effect of physical distancing measures on risk of SARS-CoV-2 transmission using empirically collected social contact data in China (10), Europe (11–13) and the United States (14) however, there is currently no such data reported for Canada. The objective of this study is to provide Canadian-specific data to evaluate the impact of physical distancing measures on the transmission of SARS-CoV-2. In doing so, we describe the age-specific contact patterns derived from survey data at four different time points during the COVID-19 pandemic, construct age-specific social contact matrices, and estimate the reproduction number for each time point.

## Methods

### Data Collection

The study protocol was approved by the University of Guelph Research Ethics Board (protocol #20-04-011) and the University of Toronto Research Ethics Board (protocol #38251). The research company, Dynata, was hired to conduct four cross-sectional electronic surveys in Canadians over the age of 18 years from May 7-19 (Survey 1), July 17-27 (Survey 2), September 21-October 10 (Survey 3), and December 8-31 (Survey 4). A quota sampling design was used and quotas for each survey were set for age, gender, official language and geographic region. Participants were recruited from a panel of survey respondents and paid a nominal amount for completing the survey. Panelists who logged into their Dynata account during the study period were directed to the survey if they fit the quotas being targeted. Survey responses were excluded from analysis if the survey was completed in less than one-third of the estimated completion time, if the respondent reported their age as less than 18 years, or if the survey was discontinued for exceeding the age, gender, or region quotas. Responses with duplicated entries for gender, age, postal code, date, and contact names were considered duplicate responses and removed from the dataset.

The survey instrument was adapted from the POLYMOD UK (6) and CoMix UK surveys (11). Respondents provided information about their age, gender, province of residence, and household composition and were then asked to record all direct contacts made between 5am on the day preceding the survey and 5am the day of survey completion including members of their household. A direct contact was defined as anyone who was met in person and with whom a short conversation occurred, or anyone with whom the respondent had physical contact (6,11). For each contact identified, respondents recorded the age of the contact and the setting in which the contact occurred. The survey instrument is available in the Appendix. As the contact diaries excluded people under the age of 18 years and with schools across the country reopening in September 2020, additional questions were added to Surveys 3 and 4. Respondents with children under 18 were asked whether any of the children in their household had attended school, taken the school bus, attended before/after school care, or participated in extracurricular activities in the 7 days prior to survey completion. Respondents were then asked to estimate the number of contacts each of their children had in each of these settings.

Given that schools had reopened, and many people were no longer working remotely by the time Survey 3 was deployed (September, 2020), respondents were also asked to identify whether their occupation required direct contact with more than 20 people during a typical work day. Respondents in these “high contact” occupations were asked to estimate the number of people in each age category with whom they would have contact at work on a typical work day. The number of reported contacts in Surveys 3 and 4 was truncated at 75 per respondent.

### Data Analysis

Respondents and contacts were categorized into age groups as follows: 18-29 years, 30-39 years, 40-49 years, 50-59 years, 60-69 years, and over 70 years. To ensure the sample was generally representative of the Canadian population, age, gender, region of residence, and household size of survey respondents were compared with the 2016 Canadian Census (15,16). Post-stratification weights were then calculated based on age and household size within Canadian region (Atlantic, Quebec, Ontario, West) using data from the 2016 Canadian census (15,16).

The average number of contacts per respondent was calculated and stratified by age group, gender, household size, region of residence, and whether the contact diary was completed for a weekday or weekend day. The average number of contacts for each survey time period was compared to the POLYMOD UK study (6), which represents pre-pandemic contacts, by calculating the percent reduction from the mean number of contacts reported in POLYMOD.

Contact matrices were constructed for the age-specific mean number of contacts per 24-hour period, adjusting for the age distribution in the Canadian population and reciprocity of contacts using the SocialMixr package in R (17). The 2016 Canadian census was used to correct for the probability of contact within the population (16). Missing contact age was sampled from other participants’ contacts within the same age group.

To provide a full contact matrix with which to estimate R_t_, contacts for the 0-4 and 5-17 year age groups were imputed using a scaled version of the POLYMOD UK (6) data by multiplying the number of contacts in corresponding age groups from the POLYMOD UK (6) study by the ratio of the dominant eigenvalues of the POLYMOD UK and the observed matrices for all age groups surveyed in both studies, stratified by location of contact (11,18). As schools were closed during the data collection periods in Surveys 1 and 2, school contacts were removed from the POLYMOD UK (6) data for the analysis of these two surveys only.

Proportions were calculated for those who reported that at least one of the children in their household participated in school-based or extracurricular activities. The average number of contacts was calculated for per child in different settings per day (school, aftercare, bus) or per week (extracurricular) among respondents reporting participation in these activities. These estimates were not included in the contact matrices.

The next generation matrix approach was used to estimate changes in the reproduction number (R_t_) (9). The reproduction number was estimated by multiplying R_0_ prior to physical distancing interventions by the ratio of the dominant eigenvalues of the POLYMOD UK (6) and observed contact matrices under the assumptions of the social contact theory that the transmission rate is proportional to rate of social contacts (5,19). A meta-analysis reported that, prior to interventions, R_0_ followed a normal distribution with a mean of 2.6 and standard deviation of 0.54 (11).

To account for sampling variability and assess uncertainty, 10,000 bootstrapped samples were generated from each of the POLYMOD UK (6) and Survey 1 – 4 contact matrices. The ratio between the dominant eigenvalues were calculated for each bootstrapped sample of the POLYMOD UK (6) and each of the observed matrices providing a distribution of the relative change in R_t_ from the observed matrices and POLYMOD UK matrices (6). This distribution was scaled with the distribution of bootstrap samples to estimate R_t_ under physical distancing measures at each of the four survey time points.

The emergence of SARS-CoV-2 variants with increased transmissibility has the potential to require more stringent public health measures (20). To assess the theoretical impact of a more transmissible variant of SARS-CoV-2, each of the scaled estimates of R_t_ were multiplied by a factor of 1.56 to provide a distribution of R_t_ estimates consistent with a 56% increase in transmissibility (20).

The sensitivity of the estimates of R_t_ to changes in child-related contacts was assessed using previously published methods (11,18). As contact diary data were collected from adults only, there is uncertainty about the average number of child-to-child and child-to-adult contacts under COVID-19 public health measures. To estimate the impact of varying the levels of child-related contacts on the estimates of R_t_, the procedure to estimate R_t_ was repeated for each Survey (1-4) with a reduction of 20%, 35%, 50%, 65%, and 80% of contacts from the POLYMOD UK study (6) for the 5-17 year age group.

All data were analysed using RStudio Version 1.2.5033 (21). The code is based on the SocialMixr package (17) as well as on the work of Jarvis et al (11). Funding to support data collection was provided by the Public Health Agency of Canada (PHAC), The National Collaborating Centre for Infectious Diseases (NCCID), and the University of Guelph. The funders had no role in study design, data collection and analysis, decision to publish or preparation of the manuscript.

## Results

We aimed to collect data from 5000 Canadians in Survey 1 (May) and 2500 Canadians in each of Surveys 2 – 4 (July, September, December). A total of 9120 survey responses were received for Survey 1, 4939 for Survey 2, 5310 for Survey 3, and 9599 for Survey 4. Respondents that completed the entire survey and were not screened out for any reason were included in the final sample resulting in 4981 responses for Survey 1, 2493 responses for Survey 2, 2495 responses for Survey 3, and 2491 responses for Survey 4. A summary of the exclusion process is shown in Appendix Figure 1.

The proportion of respondents living in each region, the male to female ratio, and the proportion of respondents in each age category were comparable to the 2016 Canadian Census of the population (Appendix Table 1).

### Contact Patterns

Descriptive statistics of reported contacts are included in Appendix Table 2. Data were analyzed for 11,019 reported contacts in Survey 1, 5608 in Survey 2, 12,289 in Survey 3, and 9703 in Survey 4 with an average number of 2.21, 2.17, 4.76, and 3.89 contacts, respectively (Table 1). These represent average reductions of 79.5%, 79.9%, 55.9%, and 64.0% for Surveys 1, 2, 3, and 4, in the number of contacts compared with pre-pandemic data from POLYMOD UK (6). The reduction in contacts was consistent across age groups in Surveys 1 and 2 however, the reduction in contacts was lower in younger age groups in Survey 3 and more variable by age in Survey 4.

**Table 1.**
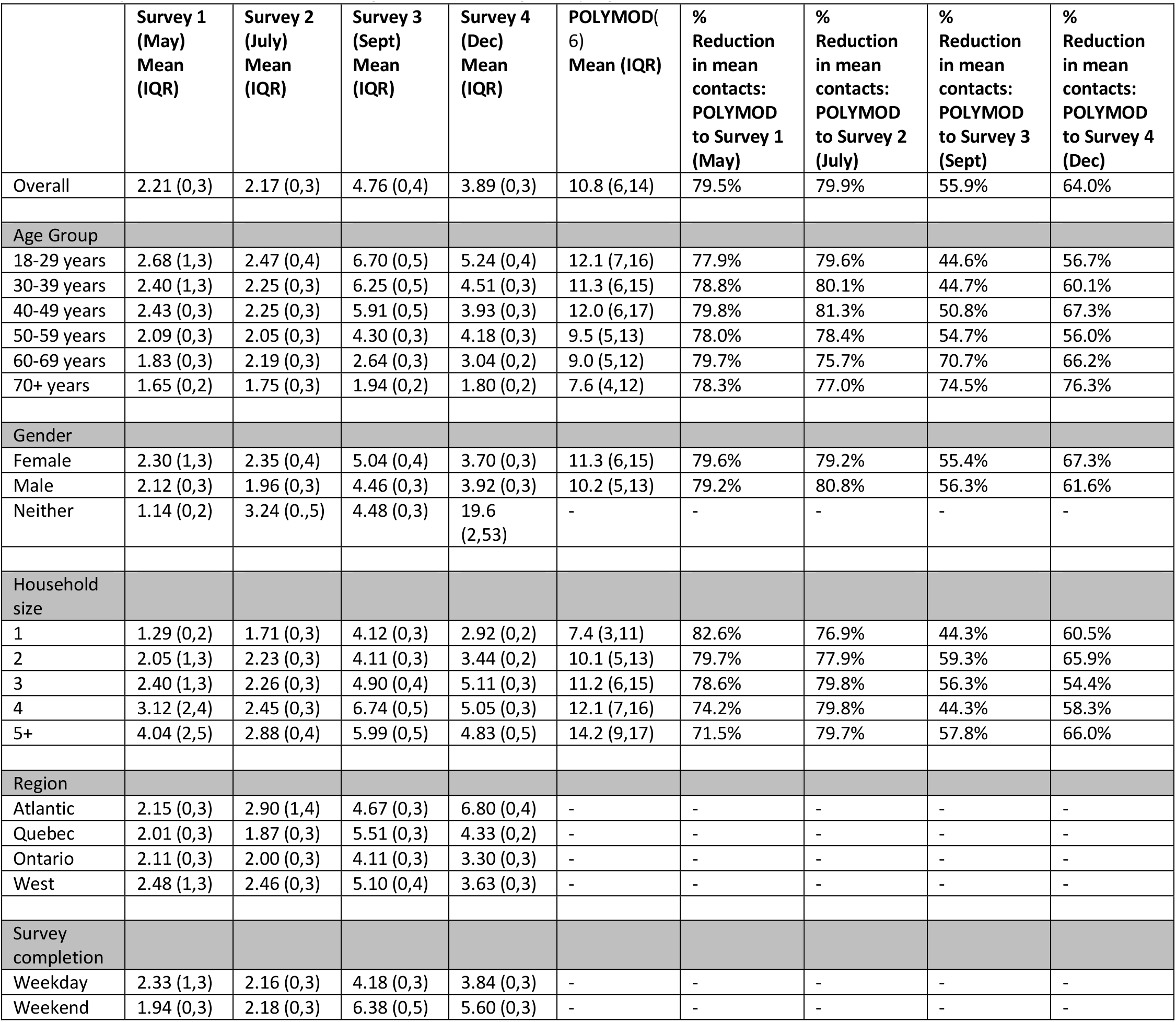
The average number of contacts for the four waves of the COVID-19 pandemic survey in Canada compared with the pre-pandemic POLYMOD UK(6) survey overall and stratified by age group, gender, household size, and whether the diary was completed for a weekday or weekend day. The average includes respondents who reported no contacts. Results are reported as mean number of contacts per 24 hour period (IQR). The total number of contacts per respondent was truncated at 75 for survey Surveys 3 and 4. Values are weighted within region by age and household size

|  | Survey 1<br>(May)<br>Mean<br>(IQR) | Survey 2<br>(July)<br>Mean<br>(IQR) | Survey 3<br>(Sept)<br>Mean<br>(IQR) | Survey 4<br>(Dec)<br>Mean<br>(IQR) | POLYMOD(6)<br>Mean (IQR) | %<br>Reduction<br>in mean<br>contacts:<br>POLYMOD<br>to Survey 1<br>(May) | %<br>Reduction<br>in mean<br>contacts:<br>POLYMOD<br>to Survey 2<br>(July) | %<br>Reduction<br>in mean<br>contacts:<br>POLYMOD<br>to Survey 3<br>(Sept) | %<br>Reduction<br>in mean<br>contacts:<br>POLYMOD<br>to Survey 4<br>(Dec) |
| --- | --- | --- | --- | --- | --- | --- | --- | --- | --- |
| Overall | 2.21 (0,3) | 2.17 (0,3) | 4.76 (0,4) | 3.89 (0,3) | 10.8 (6,14) | 79.5% | 79.9% | 55.9% | 64.0% |
| Age Group |  |  |  |  |  |  |  |  |  |
| 18-29 years | 2.68 (1,3) | 2.47 (0,4) | 6.70 (0,5) | 5.24 (0,4) | 12.1 (7,16) | 77.9% | 79.6% | 44.6% | 56.7% |
| 30-39 years | 2.40 (1,3) | 2.25 (0,3) | 6.25 (0,5) | 4.51 (0,3) | 11.3 (6,15) | 78.8% | 80.1% | 44.7% | 60.1% |
| 40-49 years | 2.43 (0,3) | 2.25 (0,3) | 5.91 (0,5) | 3.93 (0,3) | 12.0 (6,17) | 79.8% | 81.3% | 50.8% | 67.3% |
| 50-59 years | 2.09 (0,3) | 2.05 (0,3) | 4.30 (0,3) | 4.18 (0,3) | 9.5 (5,13) | 78.0% | 78.4% | 54.7% | 56.0% |
| 60-69 years | 1.83 (0,3) | 2.19 (0,3) | 2.64 (0,3) | 3.04 (0,2) | 9.0 (5,12) | 79.7% | 75.7% | 70.7% | 66.2% |
| 70+ years | 1.65 (0,2) | 1.75 (0,3) | 1.94 (0,2) | 1.80 (0,2) | 7.6 (4,12) | 78.3% | 77.0% | 74.5% | 76.3% |
| Gender |  |  |  |  |  |  |  |  |  |
| Female | 2.30 (1,3) | 2.35 (0,4) | 5.04 (0,4) | 3.70 (0,3) | 11.3 (6,15) | 79.6% | 79.2% | 55.4% | 67.3% |
| Male | 2.12 (0,3) | 1.96 (0,3) | 4.46 (0,3) | 3.92 (0,3) | 10.2 (5,13) | 79.2% | 80.8% | 56.3% | 61.6% |
| Neither | 1.14 (0,2) | 3.24 (0,5) | 4.48 (0,3) | 19.6<br>(2,53) | - | - | - | - | - |
| Household<br>size |  |  |  |  |  |  |  |  |  |
| 1 | 1.29 (0,2) | 1.71 (0,3) | 4.12 (0,3) | 2.92 (0,2) | 7.4 (3,11) | 82.6% | 76.9% | 44.3% | 60.5% |
| 2 | 2.05 (1,3) | 2.23 (0,3) | 4.11 (0,3) | 3.44 (0,2) | 10.1 (5,13) | 79.7% | 77.9% | 59.3% | 65.9% |
| 3 | 2.40 (1,3) | 2.26 (0,3) | 4.90 (0,4) | 5.11 (0,3) | 11.2 (6,15) | 78.6% | 79.8% | 56.3% | 54.4% |
| 4 | 3.12 (2,4) | 2.45 (0,3) | 6.74 (0,5) | 5.05 (0,3) | 12.1 (7,16) | 74.2% | 79.8% | 44.3% | 58.3% |
| 5+ | 4.04 (2,5) | 2.88 (0,4) | 5.99 (0,5) | 4.83 (0,5) | 14.2 (9,17) | 71.5% | 79.7% | 57.8% | 66.0% |
| Region |  |  |  |  |  |  |  |  |  |
| Atlantic | 2.15 (0,3) | 2.90 (1,4) | 4.67 (0,3) | 6.80 (0,4) | - | - | - | - | - |
| Quebec | 2.01 (0,3) | 1.87 (0,3) | 5.51 (0,3) | 4.33 (0,2) | - | - | - | - | - |
| Ontario | 2.11 (0,3) | 2.00 (0,3) | 4.11 (0,3) | 3.30 (0,3) | - | - | - | - | - |
| West | 2.48 (1,3) | 2.46 (0,3) | 5.10 (0,4) | 3.63 (0,3) | - | - | - | - | - |
| Survey<br>completion |  |  |  |  |  |  |  |  |  |
| Weekday | 2.33 (1,3) | 2.16 (0,3) | 4.18 (0,3) | 3.84 (0,3) | - | - | - | - | - |
| Weekend | 1.94 (0,3) | 2.18 (0,3) | 6.38 (0,5) | 5.60 (0,3) | - | - | - | - | - |

The age-specific contact matrices with imputed data for people younger than 18 years for each time point also reflect these reductions in the average number of contacts (Figure 1).

**Figure 1.**
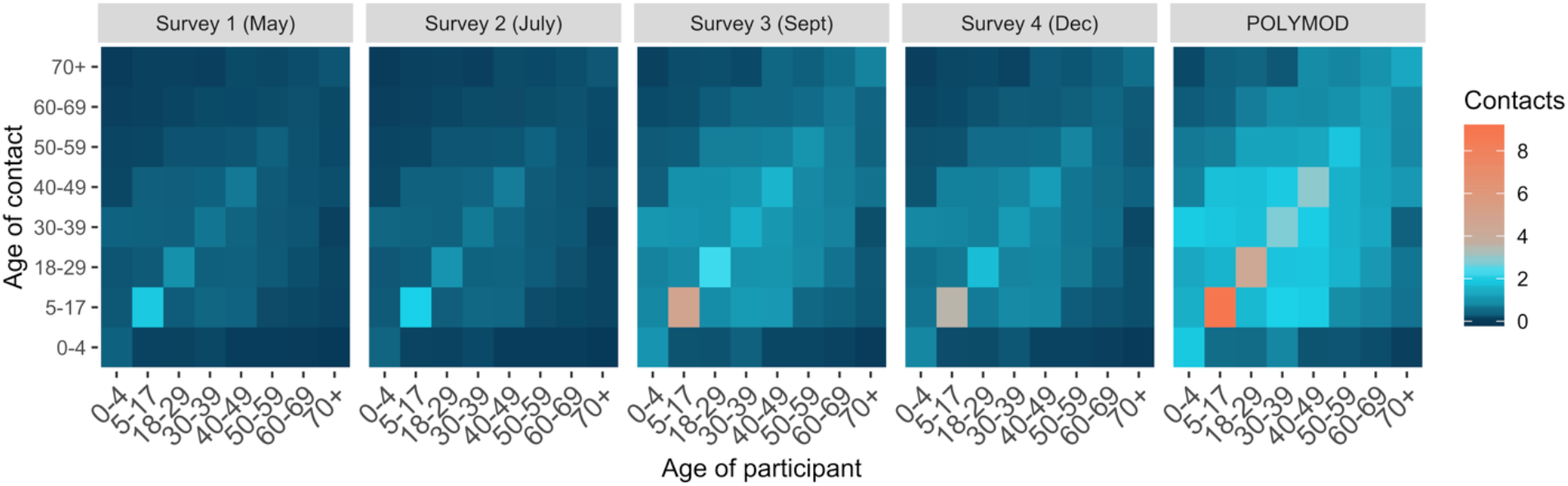
Comparison of Survey 1 (May 2020), Survey 2 (July 2020), Survey 3 (September 2020), Survey 4 (December 2020), and POLYMOD UK (6) (pre-pandemic) contact matrices. Contact matrices show the average total number of daily reported contacts by respondents in different age groups with individuals in other age groups. Child-child and child-to-adult contacts were imputed for participant age groups younger than 18 years. The number of contacts in Surveys 3 (Sept) and 4 (Dec) were truncated at 75 contacts per respondent. Data were weighted on age and household size. The 2016 Canadian census was used for demographics correcting for probability of contact within the population. Missing contact age was sampled from age-matched participants’ contacts.

### Estimates of R_t_

The estimated R_t_ values were 0.49 (95% Confidence Interval [CI]: 0.29-0.69) for Survey 1 (May), 0.48 (95% CI: 0.29-0.68) for Survey 2 (July), 1.06 (95% CI: 0.63-1.52) for Survey 3 (September), and 0.81 (0.47-1.17) for Survey 4 (December) (Figure 2). The estimated R_t_ values based on a theoretical increase in transmissibility due to the emergence of a VOC as the dominant virus strain were 0.76 (95% CI: 0.45-1.08), 0.75 (95% CI: 0.45-1.06), 1.66 (95% CI: 0.98-2.38), and 1.26 (95% CI: 0.74-1.82) for Surveys 1-4.

**Figure 2.**
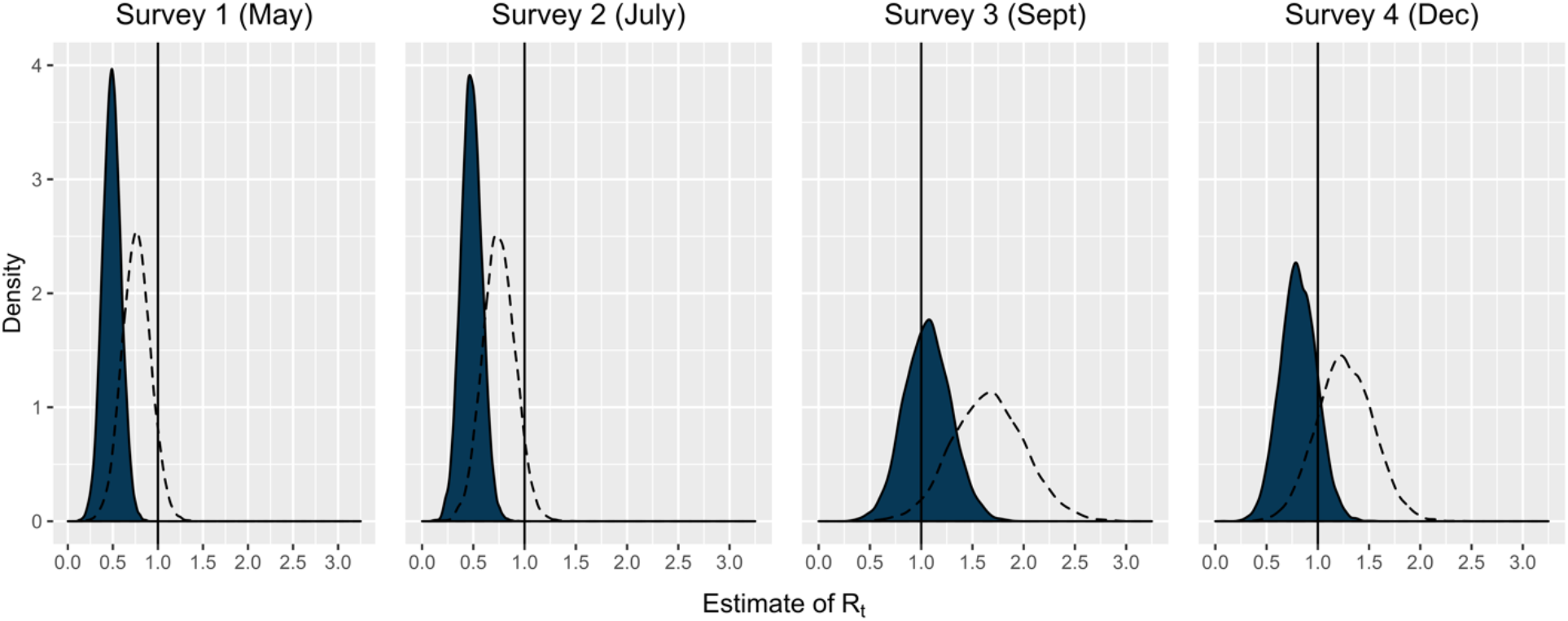
Estimated distributions of R_t_ for May, July, September, and December 2020 assuming a baseline normal distribution of R_t_ with mean = 2.6 and SD = 0.54 prior to physical distancing measures. The dotted line represents a theoretical 56% increase in transmissibility with the variant of concern VOC 202012/01 as the dominant strain (20).

The sensitivity analysis assessing the impact of reducing the number of child-related contacts by each of 20%, 35%, 50%, 65%, and 80% compared with the POLYMOD UK study (6) on estimates of Rt resulted in estimates that were similar across the different number of assumed child-related contacts at each time point (Appendix Figure 2).

The proportion of reported contacts by setting changed over the course of the study (Table 2). Respondents in Survey 1 reported the highest proportion of their contacts to have occurred within their home (51.3%) while those in Survey 2 reported the highest proportion of contacts to have occurred in ‘other’ locations such as others’ homes and community places (49.2%). Respondents in Surveys 3 and 4 reported the majority of contacts having occurred at work (66.3% and 65.4%, respectively). School contacts were largely absent from the data in all waves of the survey because contact diaries were not collected from those under the age of 18 years and most Canadian universities had transitioned to primarily remote learning for the 2020/2021 academic year.

**Table 2.** Proportion of reported contacts stratified by setting in which the contact occurred for each wave of the survey. The total number of contacts per respondent was truncated at 75 for survey Surveys 3 and 4. Results are reported as number of contacts within a 24-hour period (%).

|  | **Survey 1 (May)**<br>**n = 11,019**<br>**contacts** | **Survey 2 (July)**<br>**n = 5608**<br>**contacts** | **Survey 3 (Sept)**<br>**n = 12,289**<br>**contacts** | **Survey 4 (Dec)**<br>**n = 9703**<br>**contacts** |
| --- | --- | --- | --- | --- |
| Home | 5649 (51.3%) | 1951 (34.8%) | 1614 (13.1%) | 1384 (14.3%) |
| Workplace | 1742 (15.8%) | 855 (15.2%) | 8145 (66.3%) | 6350 (65.4%) |
| School | 52 (0.47%) | 42 (0.75%) | 140 (1.14%) | 73 (0.75%) |
| Other locations | 3576 (32.5%) | 2760 (49.2%) | 2390 (19.4%) | 1896 (19.5%) |

Figure 3 represents the average number of age-specific contacts at each time point stratified by setting in which the contact occurred. Contacts made in the respondents’ home, at school (i.e., students), and in other locations are similar across the four surveys. While the average number of workplace contacts were similar in Surveys 1 and 2, reported contacts in the workplace were notably higher in Surveys 3 and 4 (Figure 3).

**Figure 3.**
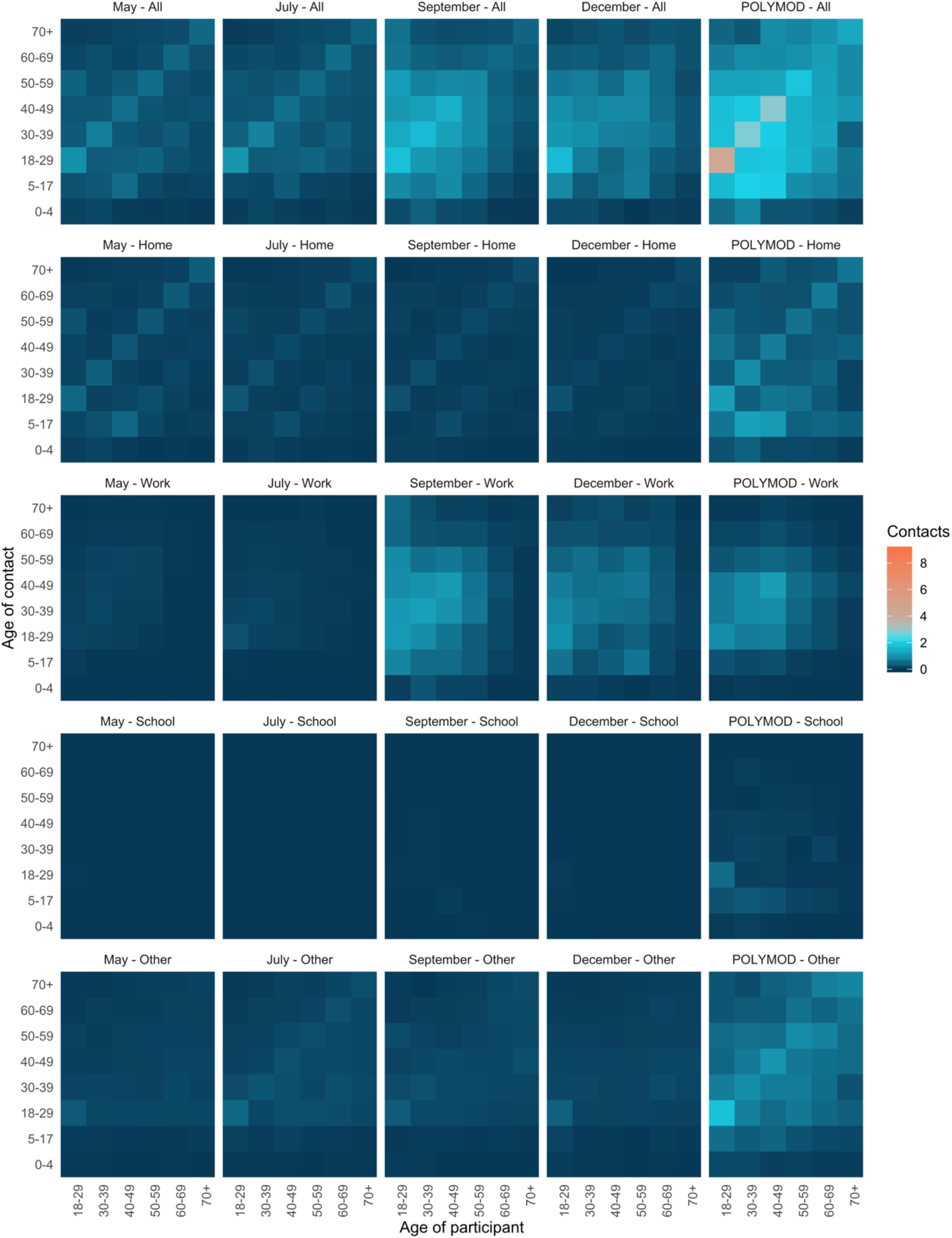
Contact matrices for all reported contacts and those stratified by setting in which the contact occurred for each wave of the survey and POLYMOD UK. Age groups younger than 18 years have been removed from the POLYMOD UK matrices to facilitate comparison with the observed matrices. Settings included all contacts, contacts made at home, contacts made in the workplace, contacts made at school, and contacts made everywhere else (including social contacts). The number of contacts in the September and December surveys were truncated at 75 contacts per respondent. Data were weighted on age and household size. The 2016 Canadian census was used for demographics in correcting for probability of contact within the population. Missing contact age was sampled from age-matched participants’ contacts.

A total of 688 (27.6%) respondents in Survey 3 and 640 (25.7%) respondents in Survey 4 reported at least one child under the age of 18 years living in their household. Of these respondents, 66.6% (95% CI: 62.9-70.0) (Survey 3) and 62.0% (95% CI 58.2-65.7) (Survey 4) reported at least one of the children in their household either participated in school-based or extracurricular activities in the 7 days prior to survey completion (Figure 4A). Respondents reported an average of 22.7 (95% CI: 21.1-24.3) (Survey 3) and 19.0 (95% CI 17.7-20.4) (Survey 4) contacts at school per day per child in attendance. For children participating in extracurricular activities, the average number of contacts reported during these activities was 18.3 (95% CI: 15.2-21.5) per week in Survey 3 and 11.2 (95% CI 8.80-13.6) per week in Survey 4 (Figure 4B).

**Figure 4A.**
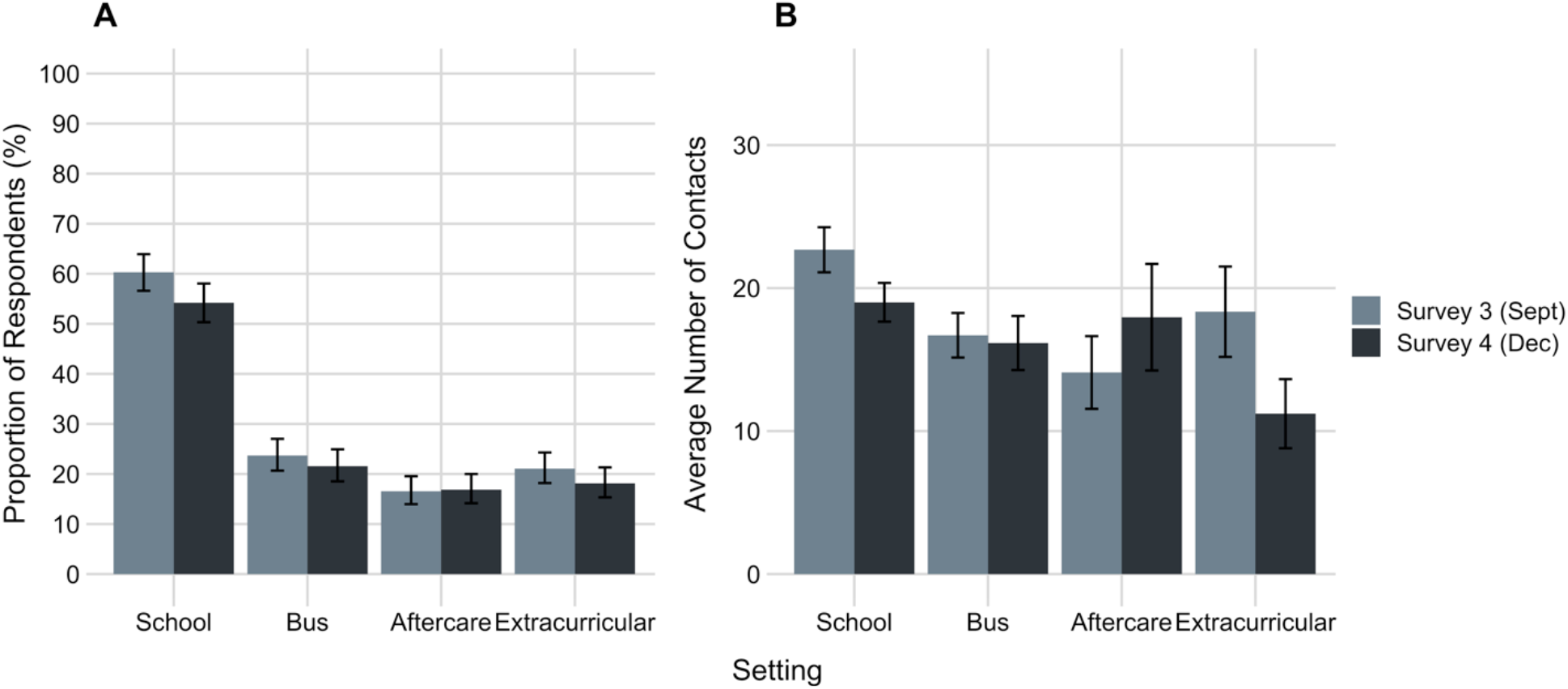
Proportion of respondents with children under the age of 18 years reporting at least one of their children attended an in-person school-based or extracurricular activity in the 7 days prior to survey completion. A total of 688 respondents in Survey 3 (Sept 2020) and 640 respondents in Survey 4 (Dec 2020) reported at least one child under the age of 18 years living in their household. **Figure 4B**. Estimated average number of contacts reported for children under the age of 18 years stratified by activity for respondents with household children who had reported that their children had participated in a school-based or extracurricular activity in the previous 7 days. Estimated contacts were reported per day (school, before/after school care, bus) or per week (extracurricular).

## Discussion

This analysis provides evidence that, in May and July (Surveys 1 and 2), Canadians were having few contacts sufficient to maintain the reproduction number well below 1, suppressing community transmission of SARS-CoV-2. The average number of contacts reported by respondents in Survey 3 (September 2020), while still more than 55% lower than those reported in the pre-pandemic POLYMOD UK study (6), had increased sufficiently to raise the reproduction number above 1 supporting a resurgence of cases of COVID-19. The contact patterns reported in Survey 4 (December 2020) likely reflect individual- and public health-level response to the epidemic growth occurring during this time period (22). The estimated R_t_ values for results for the May, July, December surveys were lower than reproduction numbers calculated using reported Canadian case data recognizing the incubation period of the virus and reporting delays (22,23) suggesting that the POLYMOD study (6) represents an overestimate of the number of pre-pandemic contacts in Canada. Indeed, while there are no published pre-pandemic contact data for Canada, a synthetic pre-pandemic contact matrix for Canada suggests that POLYMOD UK overestimates Canadian pre-pandemic contacts in many age groups (24).

According to recent modelling studies, the reduction in the average number of contacts required to suppress transmission of SARS-CoV-2 ranged from 45% to 60% (3,4,25) in comparison with study assumptions alone or with POLYMOD UK data (6), respectively. The current analysis implies epidemic growth was occurring at 55% reduction in contacts, providing empirical evidence that the reduction in contacts required to suppress a resurgence in cases is at the higher end of the spectrum of previously published estimates. Other studies of contact patterns during the COVID-19 pandemic have found that a 62-82% reduction in contacts result in R_t_ values below 1 (11,14,26) while others have found that 67-74% reduction in contacts resulted in R_t_ values greater than 1 (12,14). It must be noted that changes in R_t_ are not solely a result of changes in contact patterns but are also influenced by factors such as travel restrictions, increased use of face masks, and increased distancing when in public places (13).

The emergence of genetic variants of SARS-CoV-2 that are more easily transmitted has the potential to impact our ability to control epidemic growth. The theoretical estimates of R_t_ based on 56% higher transmissibility of the virus (20) suggest that while control measures resulting in contact patterns seen in Surveys 1 and 2 (May and July) are likely to be enough to suppress transmission, those seen in Surveys 3 and 4 (September and December) are unlikely to be sufficient to maintain R below 1 should these variants become dominant in Canada.

At the time Surveys 1 and 2 were deployed in May and July 2020, public health restrictions were beginning to be lifted and non-essential businesses and workplaces were beginning to open however, schools and daycares remained closed across the country. These restrictions are reflected in the contact matrices and associated R_t_ values in this analysis demonstrating that physical distancing was similar at the two time points. At the time of Surveys 3 and 4, daycares and schools had also reopened with the associated increase in teachers, bus drivers, and other high contact occupations returning to work. The results of our analysis are consistent with this as the higher number of contacts reported in Surveys 3 and 4 was driven by increases in contacts in the workplace rather than increases in contacts in social and other settings.

Expected increases in school contacts were not captured in this study because of the age range of respondents and the fact that many colleges and universities offered mainly remote learning opportunities at the time of the study. However, the data presented demonstrate that children are having many contacts associated with school-based and extracurricular activities. Given that large proportions of identified SARS-CoV-2 infections in children have been asymptomatic (27), these child-related contacts may represent important and overlooked opportunities for transmission. Whereas private social gatherings often bear the weight of blame for the second wave of COVID-19 cases the findings from this study suggest that contacts made during the course of the work/school day represent important opportunities for transmission. This highlights the importance of developing and enforcing stringent infection prevention and control practices in these settings.

### Limitations

The inherent risk in all surveys of being unrepresentative of the target population may have been amplified by the online nature of the survey which limited participation to those who use the Internet. Self-reporting of behaviours introduced the potential for recall and response bias. Social desirability bias carries with it the risk of underestimating the true number of total contacts as well as contacts in socially undesirable settings. These data provide no information about mitigation measures such as mask use associated with each contact. The results provide evidence that the POLYMOD UK (6) study is an imperfect representation of contact patterns in the Canadian population, however in the absence of published pre-pandemic contact data for Canada, it is a commonly used comparator. Finally, this analysis is based on contacts across Canada and does not account for any geographic variation in the number of contacts across regions potentially masking local differences.

## Conclusion

In this paper, we have quantified the effect of reducing social contact numbers on the reproduction number of COVID-19. The skewed distribution of reported contacts toward workplace settings in Waves 3 and 4 combined with the large numbers of reported school-related contacts provides evidence that these settings represent important opportunities for transmission. While transmission opportunities exist in many different settings, these data emphasize the need to support and ensure evidence-based infection prevention and control procedures in both workplaces and schools.

## Data Availability

All data and code are available from the authors upon request.

## Funding and Acknowledgements

GB and ALG are supported by the Canada Research Chairs program. DNF and ART are supported by the Canadian Institutes for Health Research (CIHR). ZP is supported by the Natural Sciences and Engineering Research Council (NSERC). EM and PJL are supported by Heritage Canada and the Social Sciences and Humanities Research Council (SSHRC). Funding to support data collection was provided by the Public Health Agency of Canada (PHAC), The National Collaborating Centre for Infectious Diseases (NCCID), and the University of Guelph. We thank J. Lau for support in programming the survey. The funders had no role in study design, data collection and analysis, decision to publish or preparation of the manuscript.

## References

1. Government of Canada. Community-based measures to mitigate the spread of coronavirus disease (COVID-19) in Canada [Internet]. 2020 [cited 2020 Dec 10]. Available from: https://www.canada.ca/en/public-health/services/diseases/2019-novel-coronavirus-infection/health-professionals/public-health-measures-mitigate-covid-19.html

2. Chief Science Officer Expert Panel on COVID-19. The Role of Bioaerosols and Indoor Ventilation in COVID-19 [Internet]. Ottawa; 2020. Available from: https://www.ic.gc.ca/eic/site/063.nsf/eng/h_98176.html

3. Tuite AR, Greer AL, De Keninck S. Risk for COVID-19 Resurgence Related to Duration and Effectiveness of Physical Distancing in Ontario, Canada. Ann Intern Med. 2020;172(1):ITC1–14.

4. Anderson S, Edwards A, Yerlanov M, Mulberry N, Stockdale J, Iyaniwura S, et al. Estimating the impact of COVID-19 control measures using a Bayesian model of physical distancing. 2020;1–15. Available from: http://dx.doi.org/10.1371/journal.pcbi.1008274

5. Wallinga J, Teunis P, Kretzschmar M. Using data on social contacts to estimate age-specific transmission parameters for respiratory-spread infectious agents. Am J Epidemiol. 2006 Nov;164(10):936–44.

6. Mossong J, Hens N, Jit M, Beutels P, Auranen K, Mikolajczyk R, et al. Social Contacts and Mixing Patterns Relevant to the Spread of Infectious Diseases. PLOS Med [Internet]. 2008;5(3):e74. Available from: https://doi.org/10.1371/journal.pmed.0050074

7. Heffernan JM, Smith RJ, Wahl LM. Perspectives on the basic reproductive ratio. J R Soc Interface. 2005;2(4):281–93.

8. Annunziato A, Asikainen T. Effective Reproduction Number Estimation from Data Series. 2020.

9. Diekmann O, Heesterbeek JAP, Roberts MG. The construction of next-generation matrices for compartmental epidemic models. J R Soc Interface. 2010;7(47):873–85.

10. Zhang J, Litvinova M, Liang Y, Wang Y, Wang W, Zhao S, et al. Changes in contact patterns shape the dynamics of the COVID-19 outbreak in China. Science (80-). 2020;368(6498):1481–6.

11. Jarvis CI, Van Zandvoort K, Gimma A, Prem K, Auzenbergs M, O’Reilly K, et al. Quantifying the impact of physical distance measures on the transmission of COVID-19 in the UK. BMC Med. 2020;18(1):124.

12. Coletti P, Wambua J, Gimma A, Willem L, Vercruysse S, Vanhoutte B, et al. CoMix: comparing mixing patterns in the Belgian population during and after lockdown. medRxiv [Internet]. 2020;2020.08.06.20169763. Available from: https://doi.org/10.1101/2020.08.06.20169763

13. Del Fava E, Cimentada J, Perrotta D, Grow A, Rampazzo F, Gil-Clavel S, et al. The differential impact of physical distancing strategies on social contacts relevant for the spread of COVID-19. medRxiv. 2020;(2).

14. Feehan D, Mahmud A. Quantifying population contact patterns in the United States during the COVID-19 pandemic. medRxiv. 2020;

15. Statistics Canada. 2016 Census. Families, households, and marital status. Statistics (Canada Catalogue no. 98-316-X2016001) [Internet]. Ottawa; 2017 [cited 2020 Nov 23]. Available from: https://www12.statcan.gc.ca/census-recensement/2016/dp-pd/prof/index.cfm?Lang=E

16. Statistics Canada. 2016 Census. Age (in Single Years) and Average Age (127) and Sex (3) for the Population of Canada, Provinces and Territories, Census Metropolitan Areas and Census Agglomerations, 2016 and 2011 Censuses. (Catalogue no. 98-400-X2016001) [Internet]. Ottawa; 2017 [cited 2020 Nov 19]. Available from: https://www12.statcan.gc.ca/census-recensement/2016/dp-pd/dt-td/Rp-eng.cfm?TABID=2&LANG=E&A=R&APATH=3&DETAIL=0&DIM=0&FL=A&FREE=0&GC=10&GL=-1&GID=1235626&GK=1&GRP=1&O=D&PID=109523&PRID=10&PTYPE=109445&S=0&SHOWALL=0&SUB=0&Temporal=2016&THEME=115&VID=0&VNAME

17. Funk S. socialmixr: Social Mixing Matrices for Infectious Disease Modelling.R package version 0.1.6. 2020.

18. Klepac P, Kucharski AJ, Conlan AJK, Kissler S, Tang M, Fry H, et al. Contacts in context: large-scale setting-specific social mixing matrices from the BBC Pandemic project. medRxiv [Internet]. 2020 Jan 1;2020.02.16.20023754. Available from: http://medrxiv.org/content/early/2020/02/19/2020.02.16.20023754.abstract

19. Hens N, Wallinga J. Design and Analysis of Social Contact Surveys Relevant for the Spread of Infectious Diseases. In: Wiley StatsRef: Statistics Reference Online [Internet]. American Cancer Society; 2019. p. 1–15. Available from: https://onlinelibrary.wiley.com/doi/abs/10.1002/9781118445112.stat07883

20. Davies NG, Abbott S, Barnard RC, Jarvis CI, Kucharski AJ, Munday JD, et al. Estimated transmissibility and impact of SARS-CoV-2 lineage B.1.1.7 in England. Science (80-). 2021;

21. RStudio Team. RStudio: Integrated Development for R. Boston, MA: RStudio, Inc.; 2019.

22. Public Health Agency of Canada. Update on COVID-19 in Canada: Epidemiology and Modelling [Internet]. 2020. Available from: https://www.canada.ca/content/dam/phac-aspc/documents/services/diseases-maladies/coronavirus-disease-covid-19/epidemiological-economic-research-data/update-covid-19-canada-epidemiology-modelling-20201211-en.pdf

23. Public Health Agency of Canada/National Microbiology Lab. COVID-19: PHAC Modelling Group Report February 25, 2021. Available from the author upon request; 2021.

24. Prem K, Cook AR, Jit M. Projecting social contact matrices in 152 countries using contact surveys and demographic data. PLoS Comput Biol [Internet]. 2017;13(9):1–21. Available from: http://dx.doi.org/10.1371/journal.pcbi.1005697

25. Tuite AR, Fisman DN, Greer AL. Mathematical modelling of COVID-19 transmission and mitigation strategies in the population of Ontario, Canada. CMAJ [Internet]. 2020; Available from: https://www.cmaj.ca/content/early/2020/04/09/cmaj.200476

26. Backer JA, Mollema L, Vos RAE, Klinkenberg D, van der Klis Frm, de Melker HE, et al. The impact of physical distancing measures against COVID-19 transmission on contacts and mixing patterns in the Netherlands: repeated cross-sectional surveys in 2016/2017, April 2020 and June 2020. medRxiv [Internet]. 2020 Jan 1;2020.05.18.20101501. Available from: http://medrxiv.org/content/early/2020/10/16/2020.05.18.20101501.abstract

27. King JA, Whitten TA, Bakal JA, McAlister FA. Symptoms associated with a positive result for a swab for SARS-CoV-2 infection among children in Alberta. Can Med Assoc J [Internet]. 2021 Jan 4;193(1):E1 LP–E9. Available from: http://www.cmaj.ca/content/193/1/E1.abstract

