## Appendix for "Quantifying Contact Patterns in Response to COVID-19 Public Health Measures in Canada"

| Exclusion process | Survey 1 | Survey 2 | Survey 3 | Survey 4 |
|---|---|---|---|---|
| Survey responses | n = 9120 | n = 4939 | n = 5310 | n = 9599 |
| Survey discontinued for exceeding quotas | n = 3960 | n = 2237 | n = 2483 | n = 6914 |
| Survey completed in less than 1/3 of estimated completion time | n = 137 | n = 187 | n = 296 | n = 162 |
| Reported age < 18 years | n = 23 | n = 15 | n = 15 | n = 23 |
| Duplicated responses | n = 19 | n = 7 | n = 10 | n =9 |
| Final sample | n = 4981 | n = 2493 | n = 2495 | n = 2491 |

**Appendix Figure 1**. Survey respondent exclusion process. Survey responses were excluded from analysis if the survey was completed in less than one-third of the estimated completion time, if the respondent reported their age as less than 18 years, or if the survey was discontinued for exceeding the age, gender, or region quotas. Responses with the duplicated entries for gender, age, postal code, date, and contact names were considered duplicate responses and removed from the dataset. Respondents that completed the entire survey and were not screened out for any reason were included in the final sample.

**Appendix Table 1.** Demographic characteristics of the sample population for Survey 1 (May 2020), Survey 2 (July 2020), Survey 3 (September 2020), and Survey 4 (December 2020) compared with demographic characteristics of the Canadian population excluding the Territories (15,16). Values are number and proportion of the whole.

| | **Number of respondents Survey 1 (May) (%)** | **Number of respondents Survey 2 (July) (%)** | **Number of respondents Survey 3 (Sept) (%)** | **Number of respondents Survey 4 (Dec) (%)** | **2016 Census (18+) (excluding Territories)** |
|---|---|---|---|---|---|
| | **n = 4981** | **n = 2493** | **n = 2495** | **n = 2491** | **N = 28,040,340** |
| **Gender** | | | | | |
| Male | 2444 (49.1%) | 1152 (46.2%) | 1162 (46.6%) | 1184 (47.5%) | 13,619,255 (48.6%) |
| Female | 2520 (50.6%) | 1327 (53.2%) | 1326 (53.1%) | 1298 (52.1%) | 14,421,165 (51.4%) |
| **Age Category** | | | | | |
| 18-29 years | 781 (15.7%) | 372 (14.9%) | 294 (11.8%) | 341 (13.7%) | 5,344,730 (19.1%) |
| 30-39 years | 935 (18.8%) | 476 (19.1%) | 497 (19.9%) | 470 (18.9%) | 4,600,440 (16.4%) |
| 40-49 years | 802 (16.1%) | 413 (16.6%) | 452 (18.1%) | 433 (17.4%) | 4,600,240 (16.4%) |
| 50-59 years | 848 (17.0%) | 507 (20.3%) | 478 (19.2%) | 474 (19.0%) | 5,283,165 (18.8%) |
| 60-69 years | 977 (19.6%) | 492 (19.7%) | 484 (19.4%) | 476 (19.1%) | 4,253,505 (15.2%) |
| 70 + years | 638 (12.8%) | 233 (9.35%) | 290 (11.6%) | 297 (11.9%) | 3,958,260 (14.1%) |
| **Region of Residence** | | | | | |
| Atlantic | 341 (6.84%) | 171 (6.86%) | 179 (7.17%) | 171 (6.86%) | 1,916,220 (6.83%) |
| Quebec | 1170 (23.5%) | 582 (23.3%) | 512 (20.5%) | 582 (23.4%) | 6,580,885 (23.5%) |
| Ontario | 1909 (38.3%) | 958 (38.4%) | 992 (39.8%) | 958 (38.5%) | 10,766,730 (38.4%) |
| Western | 1561 (31.3%) | 782 (31.4%) | 811 (32.5%) | 780 (31.3%) | 8,776,505 (31.3%) |
| **Household Size** | | | | | |
| 1 | 1154 (23.2%) | 518 (20.8%) | 564 (22.6%) | 642 (25.8%) | 3,959,395 (28.2%) |
| 2 | 1993 (40.0%) | 654 (26.2%) | 656 (26.3%) | 971 (39.0%) | 4,823,230 (34.4%) |
| 3 | 869 (17.4%) | 616 (24.7%) | 607 (24.3%) | 435 (17.5%) | 2,134,215 (15.2%) |
| 4 | 599 (12.0%) | 367 (14.7%) | 371 (14.9%) | 319 (12.8%) | 1,940,455 (13.8%) |
| 5+ | 366 (7.35%) | 338 (13.6%) | 297 (11.9%) | 124 (4.98%) | 1,174,780 (8.37%) |
| **Survey completion** | | | | | |
| Weekday | 3382 (67.9%) | 623 (25.0%) | 1873 (75.1%) | 2420 (97.1%) | - |
| Weekend | 1599 (32.1%) | 1870 (75.0%) | 621 (24.9%) | 71 (2.85%) | - |

**Appendix Table 2.** Descriptive statistics of reported contacts for each of Survey 1 (May 2020) Survey 2 (July 2020), Survey 3 (September 2020), and Survey 4 (December 2020). Cells denoted by "-" signify that the number contacts were limited to 20 entries for each of Surveys 1 and 2.

| | **Survey 1 (May) (%)** | **Survey 2 (July) (%)** | **Survey 3 (Sept) (%)** | **Survey 4 (Dec) (%)** |
|---|---|---|---|---|
| Number of respondents | 4981 | 2493 | 2495 | 2491 |
| Number of contacts | 11,019 | 5608 | 12,289 | 9703 |
| Median reported contacts per respondent | 2 | 1 | 1 | 1 |
| Range of number of contacts reported per respondent | 0-15 | 0-15 | 0-672 | 0-8524 |
| Respondent reported > 75 contacts (n (% of respondents)) | - | - | 22 (0.88%) | 25 (1.00%) |
| Median (Interquartile range) of number of contacts for > 75 reported contacts | - | - | 180 (100,340) | 130 (89,220) |
| Missing contact age (n (% of contacts)) | 0 (0%) | 6 (0.24%) | 14 (0.56%) | 125 (5.02%) |

| | | | | |
|---|---|---|---|---|
| Respondent reported no contacts (n (% of respondents)) | 1205 (24.2%) | 855 (34.3%) | 934 (37.4%) | 1118 (44.9%) |

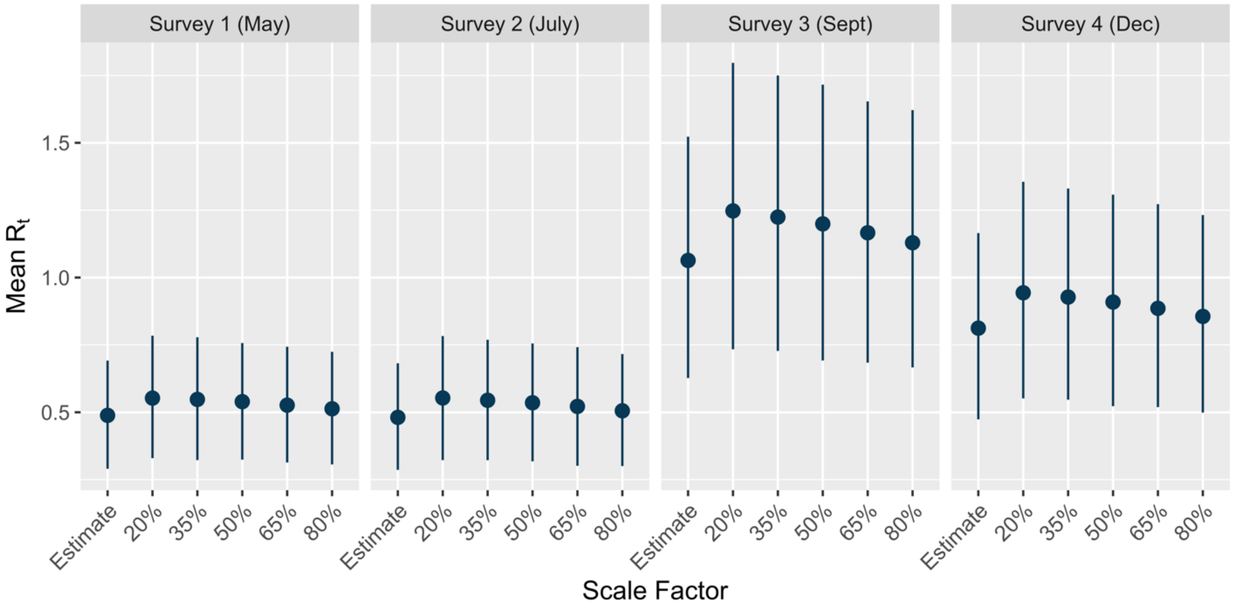


**Appendix Figure 2.** Sensitivity analysis. As contact diary data was collected from adults only, there is uncertainty about the child-to-child and child-to-adult contacts under COVID-19 public health measures. To estimate the impact of varying the levels of child-related contacts on the estimates of $R_t$, the procedure to estimate $R_t$ was repeated for each Survey (1-4) with a reduction of 20%, 35%, 50%, 65%, and 80% of contacts from the POLYMOD UK study (6) for the 5-17 year age group.

**Survey Instrument**

What is your age?

Are you...

- A man
- A woman
- Other (eg. Trans, non-binary, two-spirit, gender-queer)

Which province do you currently live in?

- Newfoundland and Labrador
- Prince Edward Island
- New Brunswick
- Nova Scotia
- Quebec
- Ontario
- Manitoba
- Saskatchewan
- Alberta
- British Columbia

What is your postal code? (Please use upper case letters only e.g. A0A 0A0)

Thinking about the place where you live, what word best describes it: A large city, a medium sized city, a large town, a small town, a rural place.

- A large city
- A medium sized city  A large town
- A small town
- A rural place

Which of the following best describes your racial or ethnic background?

- White
- Indigenous (First Nations, Inuit, Metis)
- Black
- East and Southeast Asian (e.g. Chinese, Japanese, Korean, Vietnamese, etc.)
- South Asian (e.g. Indian, Pakistani, etc.)
- West Central Asian (e.g. Iranian, Afghan)
- Middle Eastern/North African (e.g. Iraqi, Algerian)
- Pacific Islander

Can your ethnic background be described as Latin American? (i.e. Central or South American)

- Yes
- No

What was your total household income, before taxes, for the year 2018?

- No income
- $1 to $30,000
- $30,001 to $60,000
- $60,001 to $90,000
- $90,001 to $110,000
- $110,001 to $150,000
- $150,001 to $200,000
- More than $200,000
- Don't know/prefer not to answer

What is the highest level of schooling you have completed, or the highest degree you have received?

- No certificate, diploma or degree

- ○ High school diploma or equivalency certificate
- ○ Certificate of Apprenticeship or Certificate of Qualification
- ○ Other trades certificate or diploma
- ○ College, CEGEP or other non-university certificate or diploma from a program of 3 months to less than 1 year
- ○ College, CEGEP or other non-university certificate or diploma from a program of 1 year to 2 years
- ○ College, CEGEP or other non-university certificate or diploma from a program of more than 2 years
- ○ University certificate or diploma below bachelor level
- ○ Bachelor's degree
- ○ University certificate or diploma above bachelor level
- ○ Degree in medicine, dentistry, veterinary medicine or optometry
- ○ Master's degree
- ○ Earned doctorate

Which of the following best describes your current employment situation? Are you:
Employed part-time
Employed full-time
Self-employed
Unemployed
Working within the home (e.g., homemaker, stay at home parent)
Retired
Student
Other (please specify) ______________________________________________

Are you unemployed as a result of the COVID-19 pandemic?
Yes
No

What is your occupation? If retired or unemployed or on any type of leave, please indicate the category closest to your previous occupation
Management occupations
Business, finance and administration occupations

Natural and applied sciences and related occupations
Health occupations
Occupations in education, law and social, community and government services
Sales and service occupations
Trades, transport and equipment operators and related occupations
Natural resources, agriculture and related production occupations
Occupations in manufacturing and utilities
Other (please specify) __________________________________________________

How many people live in your household? Household members live at the same address and share a kitchen with you.

How many of your household members (**NOT including yourself**) fall into the following age groups? Household members live at the same address and share a kitchen with you.

Please enter 0 if there are none in an age group.
0-5 years old __________________________________________________
6-15 years old __________________________________________________
16-19 years old __________________________________________________
20-45 years old __________________________________________________
46-64 years old __________________________________________________
65-79 years old __________________________________________________
80+ years old __________________________________________________

Which of the following best describes your household?
Single person living alone
Adults only living together
Family with children living together
More than two generations living together (e.g., children, parents, and grandparents)
Skip generations living together (e.g., grandparents and grandchildren living together without the children's parents)

Is anyone in your household (**including yourself**) in a high-risk group for which the annual seasonal influenza vaccine would usually be recommended by the Public Health Agency of Canada?

(These conditions include individuals who are pregnant or those with chronic respiratory disease, chronic heart disease, chronic kidney disease, chronic liver disease, chronic neurological disease, diabetes (all types), cancer, immunosuppression, dysfunction of the spleen, and/or BMI > 40)

○ Yes

○ No

How many children do you have in your household that are under the age of 18?

- ◯ None
- ◯ 1
- ◯ 2
- ◯ 3
- ◯ 4
- ◯ 5
- ◯ 6 or more

In the **last 7 days** have any of the children in your household):

| | Yes | No | Not Applicable | Prefer not to Answer |
|---|---|---|---|---|
| Attended any type of school, daycare, or day camp | ◯ | ◯ | ◯ | ◯ |
| Taken the school bus | ◯ | ◯ | ◯ | ◯ |
| Participated in before/after school care | ◯ | ◯ | ◯ | ◯ |
| Participated in an extracurricular activity | ◯ | ◯ | ◯ | ◯ |

How many days out of the **past 7 days** have any of the children in your household:

| | 1 | 2 | 3 | 4 | 5 | 6 | 7 |
|---|---|---|---|---|---|---|---|
| Attended any type of school, daycare, or day camp | ◯ | ◯ | ◯ | ◯ | ◯ | ◯ | ◯ |
| Taken the school bus | ◯ | ◯ | ◯ | ◯ | ◯ | ◯ | ◯ |

| | | | | | | | |
|---|---|---|---|---|---|---|---|
| Participated in before/after school care | ○ | ○ | ○ | ○ | ○ | ○ | ○ |
| Participated in an extracurricular activity | ○ | ○ | ○ | ○ | ○ | ○ | ○ |

For each of the people living in your household under the age of 18, please answer the following questions:

What is the age of your child?

Please estimate the number of children and adults your child has contact with at school, daycare, or day camp in **one day** (e.g., the number of people in their classroom)

Please estimate the number of children and adults who ride the school bus with your child in **one day**

Please estimate the number of children and adults your child has contact with at before/after school care in **one day**.

Please estimate the total number of children and adults your child has contact with at their extracurricular activities in **one week**.

We will now ask you to remember and record who you had direct contact with between 5am yesterday and 5am today. We are only interested in direct contacts, which are **people who you met in person** and with whom you exchanged at least a few words, or with whom you had physical contact (e.g. a handshake, embracing, kissing).

**If you only spoke to someone over the phone or internet, they should not be included in this section.** Your contacts could include household members, friends, family, work colleagues, or people you spoke to in a store, etc.

First, did you have direct contact with anyone between 5 am yesterday and 5 am today?

○ Yes

○ No

Does your occupation require you to have **direct contact** with more than **20 people** in one work day (e.g., teacher, bus driver, etc.)?

○ Yes

○ No

Thinking about the past 7 days, please enter the total number of direct contacts you had **at work** in each of the following age groups **during one typical work day**.
Please enter 0 if there are none in an age group.

0-4 years ______________________________________________
5-17 years ______________________________________________
18-29 years ______________________________________________
30-39 years ______________________________________________
40-49 years ______________________________________________
50-59 years ______________________________________________
60-69 years ______________________________________________
70+ years ______________________________________________

Please do not report these work contacts in the next question.

Now please write a nickname for each person you had direct contact with or their description (e.g. grocery store clerk).

Note that this nickname is only needed to make it easier for you to complete the survey, so please pick a nickname that will help you identify each contact. Nicknames are not visible to anyone outside of this survey.

The order in which you give these names does not matter. However, it is easiest to give them in chronological order, e.g. when I woke up, I saw Peter and Naomi at breakfast. I then drove to my work, where I met with Jack, Deborah, and two clients. On my way back home, I stopped at a gas station, where I had a brief chat with the attendant.

Please list a nickname or description of each person you had direct contact with between 5 am yesterday and 5 am today. Enter each contact only once. Please fill in as many spaces as necessary before proceeding.

Contact [1-20]______________________________________________

What is the approximate age of [contact]?

What is [contact]'s relationship to you?

○ Member of my household

○ Someone who is part of my social contact bubble but not part of my household

○ Family member or friend who is not in my household or social contact bubble

- Someone I work with (i.e., co-worker or colleague)
- Someone who is a client or customer in my workplace
- Someone for whom I was a client or customer in their workplace
- Someone I go to school / college / university with
- Other (please specify) _______________________________________________

When you had direct contact with [contact] yesterday, did you have...?

- Physical contact (any sort of skin to skin contact e.g. handshake, embracing, or kissing)
- Non-physical contact (you did not touch the person)
- Prefer not to answer

Where did you have contact with [contact]?

- At your home
- At someone else's home
- At work
- At a place of worship
- On public transit
- At a daycare, or school
- At a store, financial institution or post office
- At a place of entertainment (e.g. restaurant)
- At a place for sports (e.g. gym)
- Outside (e.g. in a park, on the sidewalk, or street)
- Somewhere else (please specify) _______________________________________________

Please estimate the total amount of time you spent with [contact] yesterday **in minutes**.

Was this time spent indoors or outdoors?

- Indoors

○ Outdoors

○ Both

**References**


6. Mossong J, Hens N, Jit M, Beutels P, Auranen K, Mikolajczyk R, et al. Social Contacts and Mixing Patterns Relevant to the Spread of Infectious Diseases. PLOS Med [Internet]. 2008;5(3):e74. Available from: https://doi.org/10.1371/journal.pmed.0050074

15. Statistics Canada. 2016 Census. Families, households, and marital status. Statistics (Canada Catalogue no. 98-316-X2016001) [Internet]. Ottawa; 2017 [cited 2020 Nov 23]. Available from: https://www12.statcan.gc.ca/census-recensement/2016/dp-pd/prof/index.cfm?Lang=E

16. Statistics Canada. 2016 Census. Age (in Single Years) and Average Age (127) and Sex (3) for the Population of Canada, Provinces and Territories, Census Metropolitan Areas and Census Agglomerations, 2016 and 2011 Censuses. (Catalogue no. 98-400-X2016001) [Internet]. Ottawa; 2017 [cited 2020 Nov 19]. Available from: https://www12.statcan.gc.ca/census-recensement/2016/dp-pd/dt-td/Rp-eng.cfm?TABID=2&LANG=E&A=R&APATH=3&DETAIL=0&DIM=0&FL=A&FREE=0&GC=10&GL=-1&GID=1235626&GK=1&GRP=1&O=D&PID=109523&PRID=10&PTYPE=109445&S=0&SHOWALL=0&SUB=0&Temporal=2016&THEME=115&VID=0&VNAME
